# Trends in characteristics of HIV diagnosis at a tertiary HIV clinic in Nairobi, Kenya, 2010- 2020

**DOI:** 10.1101/2025.09.11.25335599

**Authors:** Jared O. Mecha, Elizabeth N. Kubo, Chemutai R.K Kenei, Collins O. Odhiambo, Kennedy Kirui, Kenneth Mutai, Albert O. Orwa, Evelyn W. Ngugi, Jonathan Mwangi

## Abstract

**Background:** Twenty percent of Kenyans aged 15-64 years did not know their HIV status in 2018. Such gaps in utilization of HIV testing by the population have impeded in-country progress in the HIV response. Understanding the characteristics of newly diagnosed HIV patients can inform the design of targeted interventions to address this gap. We describe trends in sociodemographic and clinical characteristics of individuals who were newly diagnosed with HIV at a large urban referral HIV clinic in Kenya.

**Methods:** We conducted a retrospective cohort study of people living with HIV who tested positive or presented with a positive diagnosis for the first time at the Kenyatta National Hospital (KNH) between 1^st^ January 2010 and 30^th^ September 2020. We assessed trends in sociodemographic and clinical characteristics using the Cochran-armitage test.

**Results:** We included a total of 8,595 patients in our analysis, with females accounting for 62%. A majority (91%) of patients were aged 25 years and above and were married (49%). Compared to females, males were more likely to have advanced HIV disease (AHD) [WHO stage III/IV (29% vs 36%); CD4<200 (23% vs 31%)] respectively. The proportion of new diagnoses with early disease (WHO stage I and II) increased by 10% from 2010 to 2020 (66%-76%) (p<0.001). Of all newly diagnosed persons, the proportion of 25-34 year olds increased from 6% in 2010 to 21% in 2020 (p<0.001). An upward trend was also observed among 35-44 year olds, while the proportion among children (2-8 years), adolescents (9-14 years) and youth (15-24 years) remained low and stable over the study period. HIV testing in the outpatient department was the most common testing modality in 2010 at 80% but declined to 1% in 2020 (p<0.001). The proportion of persons who were transferred to our hospital having already been tested for HIV increased from 13% to 27% (p<0.001). Fewer persons presented with current or previous opportunistic infections from 2010 to 2020.

**Conclusion:** We demonstrated gaps in timely identification of men and an increase in the proportion of those aged 25-34 years and 35-44 years among persons newly diagnosed with HIV between 2010 and 2020. We also noted an overall decline in opportunistic infections and advanced HIV disease presentations. The identified gaps suggest increasing prioritization of interventions targeted at early identification of men, and prevention of HIV infection especially in those aged 25-44 years.

## INTRODUCTION

As of 30th June 2021, an estimated 28 million of the 38 million persons living with HIV (PLHIV) worldwide were receiving life-saving antiretroviral therapy (ART) (1). Despite the remarkable progress in the HIV response, AIDS lingers on as an urgent global health crisis. With renewed global commitment to get the world on track to end HIV as a public health threat by 2030, in 2021, the Joint United Nations Programme on HIV/AIDS (UNAIDS) Programme Coordinating Board (PCB) adopted a new Global AIDS Strategy 2021-2026 (2). This bold strategy employs approaches to address gaps among populations, that have impeded the HIV response and proposes that, by 2025, the UNAIDS 95-95-95 testing and treatment targets will be achieved within all sub-populations and age groups (3).

HIV testing is the first and most critical step towards achieving the UNAIDS 95-95-95 goals. Although global HIV testing coverage was reported to be 84% in 2020 (1) substantial gaps persist. Approximately 16% of PLHIV globally and 10% of adults aged 15 years and above in sub-Saharan Africa (SSA) were unaware of their HIV status in 2020 (4). In Kenya, 20% of PLHIV aged 15-64 years were unaware of their HIV status in 2018, with the largest gaps seen in men (27%). (5). Despite the highlighted testing gaps in Kenya, there are scarce in-country data that characterizes persons newly diagnosed with HIV, especially within the context of a large, urban referral setting.

Understanding the characteristics of newly diagnosed PLHIV is critical for epidemic control (6) as it provides a lens through which targeted strategies can be designed for purposes of identifying those not yet diagnosed (7). The objective of our study was to describe trends in sociodemographic and clinical characteristics of individuals who were newly diagnosed (tested positive for the first time) with HIV or presented with a positive diagnosis for the first time at the Kenyatta National Hospital (KNH) over the period 2010 - 2020. The findings of this study will potentially address data gaps and guide further program implementation activities.

## METHODS

### Study Design

This was a retrospective cohort study of persons newly diagnosed with HIV seeking care at KNH over the period 2010-2020.

### Setting

KNH, Kenya’s largest referral and teaching hospital serving the residents of Nairobi county, peri-urban settlements and nationwide referrals, provides care to an average 80-120 HIV infected patients daily. These services are provided for free at the point of service. Patients access the KNH Comprehensive Care Centre (CCC) from various HIV testing points. These include the on-site Voluntary Counseling & Testing Centre (VCT) and provider-initiated testing and counselling (PITC) points, referrals from other facilities, and walk-ins. The range of clinical services provided at the CCC includes diagnostics for HIV and opportunistic infections (OIs), CD4 counts, ART, prophylaxis against select opportunistic infections, nutritional assessment and support, psychosocial care including HIV adherence counselling, adolescent HIV status disclosure, and supportive counselling (8). Over the study duration, funding for HIV prevention, care and treatment was largely provided by the United States President’s Emergency Plan for AIDS Relief (PEPFAR) through the University of Nairobi’s Program for Advanced HIV Care and Treatment Centers of Excellence Program (PACT-CoE, 2010 – 2015), and the Centers of Excellence in HIV Medicine Program (CoEHM, 2016-2021).

### Study Participants

All PLHIV who tested positive for the first time at KNH VCT and PITC sites during the period 1^st^ January 2010 to 30^th^ September 2020, and those who presented with a positive diagnosis for the first time and were enrolled in care at KNH during this period were eligible for inclusion. Those with missing data for most of the key variables of interest were excluded.

### Variables and measurements

Key variables of interest included: year of HIV diagnosis, testing point (onsite [VCT, PITC], offsite), sociodemographic characteristics including age, sex, residence, orphan status (for age 14 years and below), marital status (for age 18 years and above), and education level. Clinical characteristics included World Health Organization (WHO) stage at diagnosis, previous or current opportunistic infection, earliest CD4 count (within one year of diagnosis in order to limit the number of missing observations). Depending on specific variables, missing data in the threshold of 4% to 25% was categorized as ‘not documented’.

### Data management

Data were extracted from a point-of-care electronic medical record (EMR) system, IQ Care. This is an open source, browser-based system which is custom-designed for HIV care and treatment programs in resource-limited settings. Patient sociodemographic and clinical data are collected in the EMR. The routine data on key variables of interest were extracted from IQ Care for the period January 2010 to September 2020. The abstracted data were then stored in Microsoft Excel prior to analysis. Exploratory data analysis was conducted to identify outliers and inconsistent values.

All data were de-duplicated based on client unique clinic identification number and all client identifiers stripped prior to analysis. Data were stored in a password protected computer. After data extraction, the statistician did not have access to the original data in Kenya EMR and there was no way to link extracted data back to client details. The clean dataset was subsequently exported to STATA for analysis.

### Statistical analysis

We used descriptive analyses to characterize HIV positive patients at diagnosis, overall and stratified by sex for sociodemographic and clinical characteristics. Categorical variables were summarized using frequencies and proportions. Statistical comparisons were done using Chi-Square test or Fisher’s exact test as appropriate. Analysis of trends of characteristics over time was done using the Cochran-armitage test. All statistical tests were 2-sided and p-values less than 0.05 were considered significant.

### Ethics approval

Approval for these analyses was obtained from the KNH/University of Nairobi Ethics and Research Committee (P148/05/2009). Additionally, this activity was reviewed by CDC, to ensure it was conducted consistent with applicable federal law and CDC policy. § §See e.g., 45 C.F.R. part 46, 21 C.F.R. part 56; 42 U.S.C. §241(d); 5 U.S.C. §552a; 44 U.S.C. §3501 et seq.

## RESULTS

### Patient characteristics at diagnosis

A total of 8,595 patients were included in the analyses. Their characteristics at diagnosis are presented in Table 1. Females accounted for slightly over 60%, and 91% were ≥ 25 years of age (Table 1a). Most (68%) were classified as WHO stage I and II (Table 1b).

**Table 1a.** Sociodemographic characteristics of persons newly diagnosed with HIV at a tertiary HIV clinic in Nairobi, Kenya, 2010-2020.

| Characteristics | Male | Female | Overall | *p-value |
| --- | --- | --- | --- | --- |

|  | n=3300 | n=5295 | n=8595 |  |
| --- | --- | --- | --- | --- |
| Age group n (%) |  |  |  |  |
| 0-14 yrs. | 168(5) | 150(3) | 101 (4) | <0.001 |
| 15-24 yrs. | 216(7) | 203(4) | 217 (5) |  |
| 25 yrs. and above | 2916 (88) | 4942 (93) | 7858 (91) |  |
| Father alive? (for children aged 0-14 years) |  |  |  |  |
| Yes | 110 (75) | 114 (71) | 224 (73) | 0.338 |
| No | 8 (5) | 16 (10) | 24 (8) |  |
| Not documented | 28 (19) | 30 (19) | 58 (19) |  |
| Total | 146 (100) | 160 (100) | 306 (100) |  |
| Mother alive? (for children aged 0-14 years) |  |  |  |  |
| Yes | 108 (74) | 115 (72) | 224 (73) | 0.719 |
| No | 14 (10) | 20 (13) | 34 (11) |  |
| Not documented | 24 (16) | 25 (16) | 58 (19) |  |
| Total | 146 (100) | 160 (100) | 306 (100) |  |
| Marital Status n (%) (For those aged >=18 yrs.) |  |  |  |  |
| Single | 602 (20) | 1557 (31) | 2159 (26) | <0.001 |
| Married | 2015 (66) | 2020 (40) | 4035 (49) |  |
| Divorced/Separated | 209 (7) | 631 (12) | 840 (10) |  |
| Widowed | 117 (4) | 475 (9) | 592 (7) |  |
| Not documented | 123 (4) | 417 (8) | 540 (7) |  |
| Total | 3066 (100) | 5100 (100) | 8166 (100) |  |
| Education level n (%) |  |  |  |  |
| None | 87 (3) | 125 (2) | 212 (2) | 0.005 |
| Primary | 474 (14) | 826 (16) | 1300 (15) |  |
| Secondary | 677 (21) | 1067 (20) | 1744 (20) |  |
| Post-Secondary | 457 (14) | 607 (11) | 1064 (12) |  |
| Other | 10 (0) | 21 (0) | 31 (0) |  |

\*Pearson's Chi-Square

**Table 1b.** Clinical characteristics of persons newly diagnosed with HIV at a tertiary HIV clinic in Nairobi, Kenya, 2010-2020.

| **Characteristic** | **Male**<br>**n=3300** | **Female**<br>**n=5295** | **Overall**<br>**n=8595** | **\*p-value** |
| --- | --- | --- | --- | --- |
| **Year of diagnosis n (%)** |  |  |  |  |
| 2010 | 286 (9) | 469 (9) | 755 (9) | 0.258 |
| 2011 | 258 (8) | 405 (8) | 663 (8) |  |
| 2012 | 277 (8) | 412 (8) | 689 (8) |  |
| 2013 | 340 (10) | 543 (10) | 883 (10) |  |
| 2014 | 341 (10) | 552 (10) | 893 (10) |  |
| 2015 | 350 (11) | 572 (11) | 922 (11) |  |
| 2016 | 301 (9) | 594 (11) | 895 (10) |  |
| 2017 | 368 (11) | 584 (11) | 952 (11) |  |
| 2018 | 293 (8) | 450 (8) | 743 (9) |  |
| 2019 | 334 (5) | 492 (9) | 826 (10) |  |
| 2020 | 152 (4) | 222 (4) | 374 (4) |  |
| WHO Stage n (%) |  |  |  |  |
| Stage 1 & 2 | 2079 (63) | 3723 (70) | 5802 (68) | <0.001 |
| Stage 3 & 4 | 1185 (36) | 1517 (29) | 2702 (31) |  |
| Not Documented | 36 (1) | 55 (1) | 91 (1) |  |
| Earliest CD4 cell count after diagnosis n (%) |  |  |  |  |
| <200 | 1028 (31) | 1238 (23) | 2266 (26) | <0.001 |
| 200-349 | 555 (17) | 792 (15) | 1347 (16) |  |
| 350-499 | 399 (12) | 689 (13) | 1088 (13) |  |
| >500 | 520 (16) | 1263 (24) | 1783 (21) |  |
| Not Documented | 798 (24) | 1313 (25) | 2111 (25) |  |
| Median (IQR) | 342 (157-576) | 253 (110-448) | 305 (133-526) |  |
| Opportunistic Infections n (%) |  |  |  |  |
| Tuberculosis (TB) | 470 (14) | 522 (10) | 992 (12) | <0.001 |
| *Pneumocystis jirovecii* pneumonia (PCP) | 20 (1) | 21 (0) | 41 (1) | 0.198 |
| Cryptococcal Disease | 29 (1) | 24 (1) | 53 (1) | 0.016 |
| Oral Candidiasis | 57 (2) | 83 (2) | 140 (2) | 0.599 |
| Esophageal Candidiasis | 9 (0) | 15 (0) | 24 (0) | 1.000 |
| Kaposi’s Sarcoma | 12 (0) | 20 (0) | 32 (0) | 1.000 |
| Other OI* | 2313 (70) | 3551 (67) | 5844 (68) | <0.001 |
| Multiple OIs** | 2410 (73) | 3622 (68) | 6032 (70) | <0.001 |
| *Other OIs: Bacterial pneumonia, gastroenteritis (candida, cytomegalovirus, cryptosporidium), *Mycobacterium avium-intracellulare* complex), meningitis, toxoplasmosis |  |  |  |  |
| **Multiple OI: OIs listed in table + Other OIs |  |  |  |  |

\*Pearson's Chi-Square

Compared to females, males were more likely to have advanced HIV disease (AHD) [WHO stage III/IV (29% vs 36%); CD4<200 (23% vs 31%)]. The majority of the patients (70%) had a previous or current OI (Table 1b).

### Trends in patient characteristics

The changing profiles of newly diagnosed patients are presented in Table 2. While the proportion of children (2-8years), adolescents (9-14years) and youth (15-24 years) remained less than 7% during the observation period, newly diagnosed 25-34 year olds increased steadily from 6% in 2010 to 21% in 2020 (p<0.001). Similarly, the proportion of newly diagnosed 35-44 year olds increased from 28% in 2010 to 37% in 2014, then gradually fell to 32% in 2020 (p<0.001). Of note, the proportion of those aged 45 years and above decreased over the study duration.

**Table 2:** Trends in sociodemographic characteristics of newly diagnosed HIV infected persons at a tertiary HIV clinic in Nairobi, Kenya, 2010-2020.

|  | 2010 | 2011 | 2012 | 2013 | 2014 | 2015 | 2016 | 2017 | 2018 | 2019 | 2020 |  |
| --- | --- | --- | --- | --- | --- | --- | --- | --- | --- | --- | --- | --- |
|  | n=755 | n=663 | n=689 | n=883 | n=893 | n=922 | n=895 | n=952 | n=743 | n=826 | n=374 | *p-value |
| Characteristic |  |  |  |  |  |  |  |  |  |  |  |  |
| Sex n (%) |  |  |  |  |  |  |  |  |  |  |  |  |
| Male | 285 (38) | 237 (36) | 290 (42) | 343 (39) | 348 (39) | 341 (37) | 302 (34) | 371 (39) | 298 (40) | 330 (40) | 149 (40) | 0.154 |
| Female | 470 (62) | 426 (64) | 399 (58) | 540 (61) | 545 (61) | 581 (63) | 593 (66) | 581 (61) | 446 (60) | 496 (60) | 225 (60) |  |
| Age group n (%) |  |  |  |  |  |  |  |  |  |  |  |  |
| 2-4 yrs. | 0(0) | 0(0) | 0(0) | 0(0) | 0 (0) | 0 (0) | 0 (0) | 0 (0) | 0 (0) | 2 (0) | 0 (0) | <0.001 |
| 5-9 yrs. | 1(0) | 1(0) | 1(0) | 2(0) | 6 (1) | 25 (3) | 25 (3) | 27 (1) | 22 (3) | 7 (1) | 7 (2) |  |
| 10-14 yrs. | 14(2) | 21(3) | 25(4) | 27(3) | 20 (2) | 29 (3) | 24 (3) | 12 (1) | 13 (2) | 5 (1) | 3 (1) |  |
| 15-24 yrs. | 47(6) | 35(5) | 39(6) | 31(4) | 49 (5) | 39 (4) | 39 (4) | 52 (5) | 31 (4) | 39 (5) | 19 (5) |  |
| 25-34 yrs. | 44(6) | 35(5) | 42(6) | 85(10) | 86 (10) | 120 (13) | 144(16) | 169(18) | 159 (21) | 172(21) | 79 (21) |  |
| 35-44 yrs. | 215(28) | 186(28) | 196(28) | 257(29) | 328 (37) | 306 (33) | 307(34) | 337(35) | 224 (30) | 258(31) | 121 (32) |  |
| 45-54 yrs. | 259(34) | 231 (35) | 235(34) | 294(33) | 244 (27) | 253 (27) | 240(27) | 222(23) | 172 (23) | 232(28) | 97 (26) |  |
| 55+ yrs. | 175(23) | 154(23) | 151(22) | 187(21) | 160 (18) | 150 (13) | 116(13) | 133(14) | 122 (16) | 111(13) | 48 (13) |  |
| Education level n (%) |  |  |  |  |  |  |  |  |  |  |  |  |
| Primary | 7(1) | 5(1) | 4(1) | 23(3) | 49(5) | 47(5) | 35(4) | 25(3) | 21(3) | 1(0) | 0(0) | <0.001 |
| High school | 32(4) | 24(4) | 29(4) | 160(18) | 223(25) | 232(25) | 196(22) | 231(24) | 191(26) | 69(8) | 0(0) |  |
| College | 34(5) | 34(5) | 29(4) | 170(19) | 272(30) | 312(34) | 313(35) | 308(32) | 274(37) | 111(13) | 0(0) |  |
| University | 25(3) | 26(4) | 28(4) | 145(16) | 170(19) | 180(20) | 164(18) | 206(22) | 127(17) | 52(6) | 0(0) |  |
| Other | 1(0) | 0(0) | 1(0) | 2(0) | 7(1) | 2(0) | 6(1) | 10(1) | 0(0) | 3(0) | 0(0) |  |
| Not documented | 656(87) | 574(87) | 598(87) | 383(43) | 172(19) | 149(16) | 181(20) | 172(18) | 130(17) | 590(71) | 374(100) |  |
| Testing Point n (%) |  |  |  |  |  |  |  |  |  |  |  |  |
| ANC | 4(1) | 2(0) | 19(3) | 27(3) | 14(2) | 3(0) | 9(1) | 3(0) | 4(1) | 0(0) | 0(0) | <0.001 |
| IPD | 3(0) | 2(0) | 20(3) | 15(2) | 12(1) | 34(4) | 77(9) | 141(15) | 91(12) | 38(5) | 0(0) |  |
| OPD | 607(80) | 536(81) | 510(74) | 575(65) | 606(68) | 568(62) | 516(58) | 466(49) | 406(55) | 241(29) | 4(1) |  |
| Transfer in | 101(13) | 90(14) | 110(16) | 178(20) | 157(18) | 218(24) | 215(24) | 258(27) | 172(23) | 102(12) | 0(0) |  |
| Other | 6(1) | 9(1) | 17(2) | 29(3) | 44(5) | 61(7) | 43(5) | 25(3) | 22(3) | 12(1) | 1(0) |  |
| Not documented | 34(5) | 24(4) | 13(2) | 59(7) | 60(7) | 38(4) | 35(4) | 59(6) | 48(6) | § | § |  |
| WHO Stage n (%) |  |  |  |  |  |  |  |  |  |  |  |  |
| Stage 1&2 | 502(66) | 461(70) | 445(65) | 568 (64) | 623(70) | 637(69) | 579(65) | 605(64) | 471(63) | 627(76) | 284(76) | <0.001 |
| Stage 3&4 | 252(33) | 202(30) | 235(34) | 273(31) | 257(29) | 281(30) | 314(35) | 342(36) | 268(36) | 195(24) | 83(22) |  |
| Not documented | 1(0) | 0(0) | 9(1) | 42(5) | 13(1) | 4(0) | 2(0) | 5(1) | 4(1) | 4(0) | 7(2) |  |
| Earliest CD4 cell count after diagnosis n (%) |  |  |  |  |  |  |  |  |  |  |  |  |
| <200 | 208(28) | 185(28) | 241(35) | 319(36) | 291(33) | 321(35) | 266(30) | 251(26) | 98(13) | 58(7) | 28(7) | <0.001 |
| 200-349 | 171(23) | 149(22) | 136(20) | 168(19) | 173(19) | 178(19) | 139(16) | 126(13) | 59(8) | 31(4) | 17(5) |  |
| 350-499 | 151(20) | 125(19) | 108(16) | 121(14) | 136(16) | 147(16) | 108(12) | 106(11) | 51(7) | 26(3) | 9(2) |  |
| >500 | 215(28) | 191(29) | 180(26) | 226(26) | 250(25) | 226(25) | 187(21) | 145(15) | 85(11) | 65(8) | 13(3) |  |
| Not documented | 10(1) | 13(2) | 24(3) | 49(6) | 43(5) | 50(5) | 195(22) | 324(34) | 450(61) | 646(78) | 307(82) |  |
| Median (IQR) | 341(184-545) | 342.5(175-567.3) | 283(111-521) | 318(128.8-546.3) | 284.2(114.2-510) | 288.7(127.1-517.7) | 266(108.3-481.5) | 323(135-582) | 352.5(154.5-619.5) | 249(89.5-432.5) |  |  |
| Opportunistic Infections n (%) |  |  |  |  |  |  |  |  |  |  |  |  |
| TB | 114(15) | 83(13) | 112(16) | 168(19) | 101(11) | 103(11) | 98(11) | 98(10) | 63(8) | 35(4) | 17(5) | <0.001 |
| PCP | 6(1) | 7(1) | 5(1) | 3(0) | 2(0) | 0(0) | 5(1) | 6(1) | 3(0) | 3(0) | 1(0) |  |
| Cryptococcal Disease | 2(0) | 0(0) | 0(0) | 0(0) | 0(0) | 2(0) | 19(2) | 13(1) | 4(1) | 6(1) | 7(2) |  |
| Oral Candidiasis | 32(4) | 18(3) | 18(3) | 24(3) | 19(2) | 8(1) | 8(1) | 7(1) | 4(1) | 2(0) | 0(0) |  |
| Esophageal Candidiasis | 6(1) | 1(0) | 4(1) | 2(0) | 2(0) | 2(0) | 4(0) | 3(0) | 0(0) | 0(0) | 0(0) |  |
| Kaposi’s Sarcoma | 1(0) | 0(0) | 2(0) | 1(0) | 3(0) | 3(0) | 7(1) | 5(1) | 8(1) | 2(0) | 0(0) |  |
| ¥Other OI | 662(88) | 578(87) | 580(84) | 670(76) | 717(80) | 660(72) | 613(68) | 579(61) | 406(55) | 302(37) | 77(21) |  |
| ¶Any OI | 683(90) | 596(90) | 601(87) | 717(81) | 725(81) | 663(72) | 623(70) | 596(63) | 419(56) | 318(38) | 91(24) |  |

¥Other OIs: Bacterial Pneumonia, Gastroenteritis, Meningitis, Toxoplasmosis ¶Any OI: OIs listed in table + Other OIs
\*Cochran-armitage test
§ Data not displayed as over 50% of 2019 data and 99% of 2020 did not document this variable

There was a steep decline in the proportion of newly diagnosed patients tested in the outpatient department (OPD) (from 80% in 2010 to 29% in 2019 (p<0.001). Conversely, the proportion of those newly diagnosed in the inpatient department (IPD) progressively increased from zero in 2010 to 15% in 2017, then declined to 5% in 2019 (p<0.001). Transfers-in increased from 13% in 2010 to 27% in 2017.

The proportion with early HIV disease (WHO stage I and II) increased by 10% from 2010 to 2020 (p<0.001) while advanced disease and CD4 <200 declined from 33% in 2010 to 22% in 2020, and 28% in 2010 to 7% in 2020 respectively (p <0.001) (Table 2). Declining trends were also observed for OIs. (P <0.001) (Table 2).

## Discussion

This ten year analyses of close to 9,000 patients at Kenya’s largest tertiary healthcare institution highlights the changing characteristics of newly diagnosed PLHIV between 2010 and 2020. First, while the proportion of young adults and middle aged persons living with HIV increased over time, that of children, adolescents and youth remained low and stable, while that of 44 year olds and above declined. Second, there was a steep decline in the proportion of new HIV diagnoses in the outpatient testing points and an increase in the inpatient department as well as transfers-in. Third, there was an increased tendency for early HIV WHO clinical Stages 1 and 2.

The majority of newly diagnosed persons during the 10 year observation period were female. This is a reflection of the national sex distribution of HIV, where double the proportion of women compared to men were HIV infected (5). Findings from a national population survey also indicate that women are more likely to be aware of their HIV status than men (5). The female predominance could also be a result of fewer men accessing HIV testing services, with lower proportions of them enrolling into HIV care and initiating ART (9, 10). Women’s contact with the health system during pregnancy and child birth also contributes to the difference (11). Consistent with previous in-country studies (9, 12), most of the newly diagnosed HIV-infected persons were married. Similar findings have also been documented in other sub-Saharan African countries (13), a reflection of the generalized nature of the HIV epidemic (14) in this region.

The observed increase in new diagnoses among young adults and middle aged persons is a reflection of the national age distribution of HIV (5) as well as increased awareness of testing due to countrywide mass media campaigns (12). Our study reported low but stable proportions of new diagnoses among children aged less than five years. This was in contrast to the findings of a Central Kenya study which documented a decline from 8.7% in 2004 to 1.1% in 2014 in this age group. Both observations are attributable to effective prevention of mother-to-child transmission (PMTCT) interventions, noting that the KNH program was among the earliest to attain elimination of mother-to-child transmission (eMTCT) in the country. These findings are consistent with global program data reports of a 54% decline in new HIV infections in children from 2010 to 2020 due to increased coverage of PMTCT (15). Comparable to the observation made in the under fives, the 5-9 years and 10-14 year olds’ new diagnoses remained consistently low during the observation period. Worryingly, this may be suggestive of low testing coverage in these populations. Recent UNAIDS reports indicate that close to two thirds of children who are not on treatment fall in the 5-14 year age group and they recommend prioritization of index, family and household testing to aid in diagnosis and close the treatment gap (15).

Adolescents and young people in Kenya contribute to approximately 30% of all new HIV infections, making them a priority population for HIV interventions (16). However, our study demonstrated consistently low proportions of new diagnoses among 10-24 year olds, suggestive of gaps in coverage of HIV testing. Analyses of data from 16 SSA countries established that adolescents and young people (AYPs) had the lowest odds of testing compared to other age groups (11). Barriers to testing in this population include fear of disclosure (17) and laws restricting access to testing or requiring parental consent (18). Established strategies to address the testing gap include increasing community based testing, integrating HIV testing with other health services such as reproductive health, and developing differentiated service models (17, 19). Community-based delivery of oral HIV self-testing with individualized ongoing support during and after testing may expand access to testing among AYP (20).

While the majority of new HIV diagnoses were identified in the OPD, new diagnoses in the OPD declined during the 10 year observation period. This could have been partly due to a shift in internal policies that required very sick patients to be tested in the IPD as opposed to the OPD. Additionally, due to the referral nature of the facility, a proportion of the patients received have already been diagnosed elsewhere. Furthermore, low patient turnout during the COVID-19 pandemic possibly played a role.

We noted a progressive increase in the proportion of patients diagnosed in early clinical stages of HIV and with improved immune status. The 2008 national scale-up of Provider Initiated Testing and Counselling (PITC) (21), and implementation of the Universal Test and Treat (UTT) (22, 23) strategy after 2015 are possible explanations for this observation. In addition, better testing strategies such as index testing could have played a role. A study done in Rwanda documented a 36.8% decline in the proportion enrolling in HIV care and initiating ART with advanced HIV disease from 2007 to 2012, following national ART eligibility guideline changes in 2007-2008. The same analyses also reported increased median CD4 count from 183 cells/ µl to 293 cells/µl over the 5 year observation period (24).

Our study showed a declining trend in the proportion of OIs among newly diagnosed persons with HIV. This is consistent with the observed increase in the proportion of early diagnoses. Specifically, we observed a declining trend in previous or current TB from 15% to 5%. A 2016 national TB prevalence survey in the general adult population reported a prevalence of 558 per 100,000, with less than 50% reporting cough>2weeks (30). Considering that the prevalence of TB is expected to be higher among those immunocompromised, it is likely that our findings are an underestimate of the actual proportion of TB in our patient cohort. The national TB survey reported the highest disease burden among 25-34 year olds and males (30), both being populations where we noted gaps in HIV identification, possibly contributing to our relatively low rates.

The majority of late stage diagnoses were among men. A 2015-2020 study in Mozambique identified male sex as one of the risk factors for late presentation (AOR=2.41) (25). Similar findings have previously been documented in Ethiopia (26), South Africa (27), and Benin (28). Previous work done in Kenya identified a number of barriers to male HIV testing, including perceived providers’ attitudes, facility location and set up, wait time/inconvenient clinic times, low perception of risk, limited HIV knowledge, stigma, discrimination and fear of having a test (29). Suggested interventions targeting men include the distribution of self test kits to pregnant and postpartum women to take home to their spouses, home based testing, and involving men in the antenatal clinic (ANC) setup (11), during which they get tested.

Strengths of this study include the overall large sample size which served to provide reasonable power for precision in measurement. The ten year observation period allowed for trend analysis. Limitations of the study include the retrospective nature of the study with resultant data quality challenges including missing information especially from 2018 onwards. Another limitation is the cross sectional nature of the study in which temporality cannot be established hence we cannot infer causal associations.

## Conclusions

Our study demonstrated gaps in timely identification of men and an increase in the proportion of newly HIV diagnosed among those aged 25-34 years and 35-44 years, compared with other age categories. We also noted an overall decline in advanced HIV disease presentations. The identified gaps call for prioritization of interventions targeted at early identification of HIV infected men, and prevention of HIV infection especially in those aged 25-44 years.

## Data Availability

The data supporting this study are not publicly available due to restrictions imposed by Kenyatta National Hospital (KNH) policy. However, data may be obtained from the KNH Institutional Data Access/Ethics Committee for researchers who meet the criteria for access to confidential information. Requests can be directed to the Committee Chair. Ethical approval for this study was granted by the KNH/University of Nairobi Ethics and Research Committee (Ref: P148/05/2009).

## Funding Attribution

This project was supported by the President’s Emergency Plan for AIDS Relief (PEPFAR) through the US Centers for Disease Control and Prevention (CDC) under the terms of the Cooperative Agreement Number #U2GPS002182-01-05

## Disclaimer

The findings and conclusions in this manuscript are those of the author(s) and do not necessarily represent the official position of the funding agencies.

## Conflict of Interest Declaration

The authors disclose no conflicts of interest.

## Notes

### Competing Interest Statement

The authors have declared no competing interest.

### Funding Statement

Yes

### Author Declarations

KNH/University of Nairobi Ethics and Research Committee (P148/05/2009). Additionally, this activity was reviewed by CDC, to ensure it was conducted consistent with applicable federal law and CDC policy.

## List of References

1. UNAIDS. Global HIV & AIDS statistics - 2021 Fact sheet Geneva: UNAIDS; 2022 [cited 2022 4th April 2022]. Available from: https://www.unaids.org/en/resources/fact-sheet.

2. UNAIDS. End Inequalities. End AIDS. Global AIDS Strategy 2021-2026 Geneva: UNAIDS; 2022 [cited 2022 4th April, 2022]. Available from: https://www.unaids.org/en/Global-AIDS-Strategy-2021-2026.

3. UNAIDS. Global AIDS Strategy 2021-2026. End Inequalities. End AIDS. Geneva: 2022.

4. UNAIDS. 2021 UNAIDS Global AIDS Update - Confronting inequalities - Lessons for pandemic responses from 40 years of AIDS Geneva: UNAIDS; 2022 [cited 2022 6th April, 2022]. Available from: https://www.unaids.org/en/resources/documents/2021/2021-global-aids-update.

5. National AIDS and STI Control Programme (NASCOP). Preliminary KENPHIA 2018 Report. Nairobi, Kenya: NASCOP; 2020.

6. R. Harklerode, W. Waruiru, F. Humwa, A. Waruru, T. Kellogg, L. Muthoni, et al. Epidemiological profile of individuals diagnosed with HIV: results from the preliminary phase of case-based surveillance in Kenya. AIDS care. 2020;32(1):43–9.

7. Consolidated guidelines on person-centred HIV patient monitoring and case surveillance [Internet]. World Health Organization. 2017. Available from: https://www.who.int/publications/i/item/978-92-4-151263-3.

8. Jared O. Mecha, Elizabeth N. Kubo, Lucy W. Nganga, Peter N. Muiruri, Lilian N. Njagi, Immaculate N. Mutisya, et al. Trends in clinical characteristics and outcomes of Pre-ART care at a large HIV clinic in Nairobi, Kenya: a retrospective cohort study. AIDS Res Ther. 2016;13:38–.

9. P. Wekesa, A. McLigeyo, K. Owuor, J. Mwangi, L. Isavwa, A. Katana. Temporal trends in pre-ART patient characteristics and outcomes before the test and treat era in Central Kenya. BMC Infect Dis. 2021;21(1):1007.

10. Paul Wekesa, Angela McLigeyo, Kevin Owuor, Jonathan Mwangi, Evelyne Nganga, Kenneth Masamaro. Factors associated with 36-month loss to follow-up and mortality outcomes among HIV-infected adults on antiretroviral therapy in Central Kenya. BMC Public Health. 2020;20(1):328.

11. S. Staveteig, T. N. Croft, K. T. Kampa, S. K. Head. Reaching the ‘first 90’: Gaps in coverage of HIV testing among people living with HIV in 16 African countries. PloS one. 2017;12(10):e0186316.

12. J. O. Mecha, E. N. Kubo, L. W. Nganga, P. N. Muiruri, L. N. Njagi, S. Ilovi, et al. Trends, treatment outcomes, and determinants for attrition among adult patients in care at a large tertiary HIV clinic in Nairobi, Kenya: a 2004-2015 retrospective cohort study. HIV/AIDS (Auckland, NZ). 2018;10:103–14.

13. Nurilign Abebe Moges, Olubukola Adeponle Adesina, Micheal A. Okunlola, Yemane Berhane. Same-day antiretroviral treatment (ART) initiation and associated factors among HIV positive people in Northwest Ethiopia: baseline characteristics of prospective cohort. Archives of Public Health. 2020;78(1):87.

14. B. Kharsany, Q. A. Karim. HIV Infection and AIDS in Sub-Saharan Africa: Current Status, Challenges and Opportunities. The open AIDS journal. 2016;10:34–48.

15. UNAIDS Data 2021. Programme on HIV/AIDS [Internet]. 2021. Available from: https://www.unaids.org/sites/default/files/media_asset/JC3032_AIDS_Data_book_2021_En.pdf.

16. National AIDS Control Council (NACC). Kenya’s Fast-track plan to end HIV and AIDS among Adolescents and Young People. Internet 2015.

17. Elizabeth Glaser Paediatric AIDS Foundation (EGPAF). Adolescents and HIV: Prioritization for Elizabeth Glaser Pediatric AIDS Foundation Programs, Advocacy and Research. Washington, DC: 2017.

18. UNICEF. For Every Child, End AIDS-Seventh Stocktaking Report. New York, NY: UNICEF; 2016.

19. WHO. HIV and adolescents: guidance for HIV testing and counselling and care for adolescents living with HIV: Recommendations for a public health approach and considerations for policy-makers and managers. Geneva, Switzerland: World Health Organization; 2013.

20. R. Lapsley, K. Beima-Sofie, H. Moraa, V. Manyeki, C. Mung’ala, P. K. Kohler, et al. “They have given you the morale and confidence:” adolescents and young adults want more community-based oral HIV self-testing options in Kenya. AIDS care. 2023;35(3):392–8.

21. National AIDS and STI Control Programme (NASCOP). Guidelines for HIV testing and counselling in Kenya. In: sanitation Mopha, editor. 2010.

22. R. M. Granich, C. F. Gilks, C. Dye, K. M. De Cock, B. G. Williams. Universal voluntary HIV testing with immediate antiretroviral therapy as a strategy for elimination of HIV transmission: a mathematical model. Lancet (London, England). 2009;373(9657):48–57.

23. Ministry of Health. Guidelines on the use of antiretroviral drugs for treating and preventing HIV infection in Kenya-2016 edition. In: Programme NAaSC, editor. 2016.

24. E. Mutimura, D. Addison, K. Anastos, D. Hoover, J. C. Dusingize, B. Karenzie, et al. Trends in and correlates of CD4+ cell count at antiretroviral therapy initiation after changes in national ART guidelines in Rwanda. AIDS (London, England). 2015;29(1):67–76.

25. J. S. Chone, A. B. Abecasis, L. Varandas. Determinants of Late HIV Presentation at Ndlavela Health Center in Mozambique. International journal of environmental research and public health. 2022;19(8).

26. H. Belay, F. Alemseged, T. Angesom, S. Hintsa, M. Abay. Effect of late HIV diagnosis on HIV-related mortality among adults in general hospitals of Central Zone Tigray, northern Ethiopia: a retrospective cohort study. HIV/AIDS (Auckland, NZ). 2017;9:187–92.

27. P. K. Drain, E. Losina, G. Parker, J. Giddy, D. Ross, J. N. Katz, et al. Risk factors for late-stage HIV disease presentation at initial HIV diagnosis in Durban, South Africa. PloS one. 2013;8(1):e55305.

28. Djimon Marcel Zannou, Pacos Bray Gandaho, Angèle Azon-Kouanou, Carin Ahouada, Kuessi Anthelme Agbodande, Armand Wanvoegbe, et al. Late presentation to care among people living with HIV in Cotonou, Benin: a retrospective analysis from 2003 to 2014. Open Journal of Internal Medicine. 2017;7(4):123–34.

29. J. Okal, D. Lango, J. Matheka, F. Obare, C. Ngunu-Gituathi, M. Mugambi, et al. “It is always better for a man to know his HIV status” - A qualitative study exploring the context, barriers and facilitators of HIV testing among men in Nairobi, Kenya. PloS one. 2020;15(4):e0231645.

30. M. Enos, J. Sitienei, J. Ong’ang’o, B. Mungai, M. Kamene, J. Wambugu, et al. Kenya tuberculosis prevalence survey 2016: Challenges and opportunities of ending TB in Kenya. PloS one. 2018;13(12):e0209098.

